# Is the Healthy Start scheme associated with increased food expenditure in low-income families with young children in the United Kingdom?

**DOI:** 10.1101/2020.11.04.20225094

**Authors:** Jennie Parnham, Christopher Millett, Kiara Chang, Anthony A Laverty, Stephanie von Hinke, Jonathan Pearson-Stuttard, Eszter P Vamos

## Abstract

**Introduction:** Healthy Start is a food assistance programme in the United Kingdom (UK) which aims to enable low-income families on welfare benefits to access a healthier diet through the provision of food vouchers. Healthy Start was launched in 2006 but remains under-evaluated. This study aims to determine whether participation in the Healthy Start scheme is associated with differences in food expenditure in a nationally representative sample of households in the UK.

**Methods:** Cross-sectional analyses of the Living Costs and Food Survey dataset (2010-2017). All households with a child (0-3 years) or pregnant woman were included in the analysis (n=4,869). Multivariable quantile regression compared the expenditure and quantity of fruit and vegetables (FV), infant formula and total food purchases. Four exposure groups were defined based on eligibility, participation and income (Healthy Start Participating, Eligible Non-participating, Nearly Eligible low-income and Ineligible high-income households).

**Results:** Of 876 eligible households, 54% participated in Healthy Start. No significant differences were found in FV or total food purchases between participating and eligible non-participating households, but infant formula purchases were lower in Healthy Start participating households. Ineligible higher-income households had higher purchases of FV.

**Conclusion:** This study did not find evidence of an association between Healthy Start participation and FV expenditure. Moreover, inequalities in FV purchasing persist in the UK. Higher participation and increased voucher value may be needed to improve programme performance and counteract the harmful effects of poverty on diet.

## BACKGROUND

Individuals in the UK from higher socioeconomic positions are 80% more likely than those from lower socioeconomic positions to eat the recommended amount of fruit and vegetables (FV) a day(1). As diet is one of the leading risk factors for non-communicable disease morbidity and mortality, inequalities in dietary intake contribute to health inequalities in the UK(2). The cost of a healthy diet is proposed as a key factor among the complex and multifaceted determinants of socioeconomic inequality in diets(3–5). Moreover in recent years, the price of FV has increased disproportionately relative to nutrient-poor, energy-dense foods(6, 7), serving to exacerbate the financial barrier to a healthy diet.

The UK’s Healthy Start scheme was introduced in 2006 with the aim of improving the access to a healthy diet for low-income families(8). The scheme entitles low-income families with a pregnant woman or child aged 0-3 years to receive vouchers which can be redeemed for FV, cow’s milk and infant formula. The vouchers are worth £3.10/week per pregnant woman and child aged 1-3 years and £6.20/week per child under one year. Access to the scheme is not automatic, beneficiaries must apply. As such, uptake is strongly dependant on health professionals signposting possible participants in pre- and post-natal healthcare appointments(9).

The impact of Healthy Start is under-studied and therefore, not well-understood. Qualitative evaluations have found that Healthy Start vouchers were valued by recipients and helped reduce the impact of food insecurity(9). However, the only two existing large-scale quantitative evaluations of Healthy Start are in contradiction; reporting a null effect on FV intake(10) and positive effect on FV purchasing(11), respectively. These two previous evaluations used eligibility, not participation, as the exposure variable. Not all eligible households participate in Healthy Start, with evidence suggesting that programme uptake has been falling in recent years(12). It is currently unknown which household characteristics are associated with participation in Healthy Start. Moreover, there is no evidence on whether participation is associated with different spending within the eligible population. It is important for policy makers to understand if Healthy Start reaches its target population and whether it is effective at improving the nutrition of low-income families.

This paper aims to determine whether Healthy Start participation is associated with differences in purchasing of FV, infant formula and total food purchases among households who are Healthy Start participants, eligible non-participants, nearly eligible non-participants and ineligible non-participants.

## METHODS

### Data source and study participants

The Living Costs and Food Survey (LCFS) is an annual cross-sectional survey of UK households which collects detailed income, expenditure and sociodemographic data(13). The sample is a multi-stage stratified random sample with clustering, selected from a register of postcodes in the UK(14). Full details of survey methodology are reported elsewhere(14). Participating households were surveyed at home by a trained interviewer and instructed to complete a two-week expenditure diary collecting the expenditure and quantity of purchases. LCFS measured Healthy Start participation from 2010 onwards. Due to low annual sample size of participating households, a cross-sectional study design was used, pooling years 2010-2017.

### Analytic sample

All households with a pregnant woman or child 0-3 years were included in the analytic sample, in congruence with the Healthy Start eligibility criteria (see Table S1). Healthy Start vouchers are dispensed at the household level, so the household was used as the unit of analysis. There was a total of 42,034 households surveyed across years 2010-2017 in the LCFS. Households without a child 0-3 years or pregnant woman (*n*=37,147) and households with missing data (*n*=25) were excluded, leaving 4,869 households in the study.

### Exposure groups

Data on income and welfare benefits were collected through interview and confirmed through official documentation (e.g. payslips). Income was equivalised using OECD scales to account for the effect of household size and composition on expenditure(14). All the households in the study sample had a household member who would qualify for Healthy Start by their age or pregnancy status, thus income-level was used to stratify the exposure groups.

Households were categorised as eligible for Healthy Start if they received a qualifying income-related welfare benefit (see Table S1). This group was further divided by participation in the Healthy Start scheme. The remaining households did not receive a qualifying benefit, therefore were ineligible for Healthy Start. Ineligible households were also divided into two groups, low- and high-income households. Households were defined as low-income if they had an income less than 60% of the median disposable income that year, after adjustment for inflation(15). The low-income group represented households who just missed out on welfare schemes but were still at a high risk of experiencing food insecurity. The high-income group was included to explore and quantify differences in household expenditure across the socioeconomic gradient.

In summary, the four exposure groups derived were:

1. *Healthy Start Eligible Participants (Group [G]1)*; households who received an income-related welfare benefit and reported receiving Healthy Start vouchers.
2. *Healthy Start Eligible Non-participants (G2)*; households who received an income-related welfare benefit but did not report receiving Healthy Start vouchers.
3. *Nearly Eligible Non-participants (G3);* households defined as low-income but did not receive benefits.
4. *Ineligible Non-participant*s (*G4*); households who were neither low-income nor received benefits.

### Outcome variables

Expenditure and quantity of all purchases were recorded in a two-week expenditure diary, confirmed by receipts. Analysis using quantity variables as outcomes were restricted to years 2010-2015 as later data on quantity of purchases have not yet been released. Both the expenditure and quantity of food purchases were used as outcomes to explore whether households chose different priced products within the same category. For example, low-income households are more likely to be price sensitive(16), and therefore may choose fruit and vegetables which are lower in cost, but not volume. This is important as only differences in the quantity of food purchased have important implications for health.

The following variables were used as outcome variables, all averaged across one week per household: (i) FV expenditure (£/week); (ii) Healthy Start qualifying foods (fresh or frozen FV, cow’s milk and infant formula) expenditure (£/week); (iii) infant milk expenditure (£/week); (iv) total food expenditure (£/week); (v) FV quantity (kg/week) and (vi) Healthy Start foods quantity (kg/week).

### Covariates

Covariates included survey year, survey quarter, household size, number of children in the household, age of household reference person (HRP)(years), ethnicity of HRP (White or Black and Asian Minority Ethnicities [BAME]), National Statistics Socioeconomic Classification (NS-SEC) social class (higher professional occupations, intermediate occupations, routine and manual occupations and unemployed or students), age HRP completed full-time education (<16 years; 16-18 years and >18 years) and region (North, Midlands, East, London, South, Wales, Scotland and Northern Ireland).

### Statistical analysis

To account for inflation, income and expenditure variables were adjusted using category specific Consumer Price Indices, using 2017 as the base year(17). Indicators for survey year and quarter were included to control for macroeconomic differences across time. Survey weights, generated by LCFS, were used in all analyses to account for non-response bias and to produce results representative to the population(14). Analyses using infant formula as an outcome were performed on a subsample of households with a child less than one-year old (*n*=1,260), as the vouchers may only be redeemed for infant milk for this age-range.

We used appropriate significance tests to examine the characteristics between (i) Healthy Start Eligible Participants (G1) and Eligible Non-participants (G2); and (ii) across all four exposure groups (See Table 1).

**Table 1.**
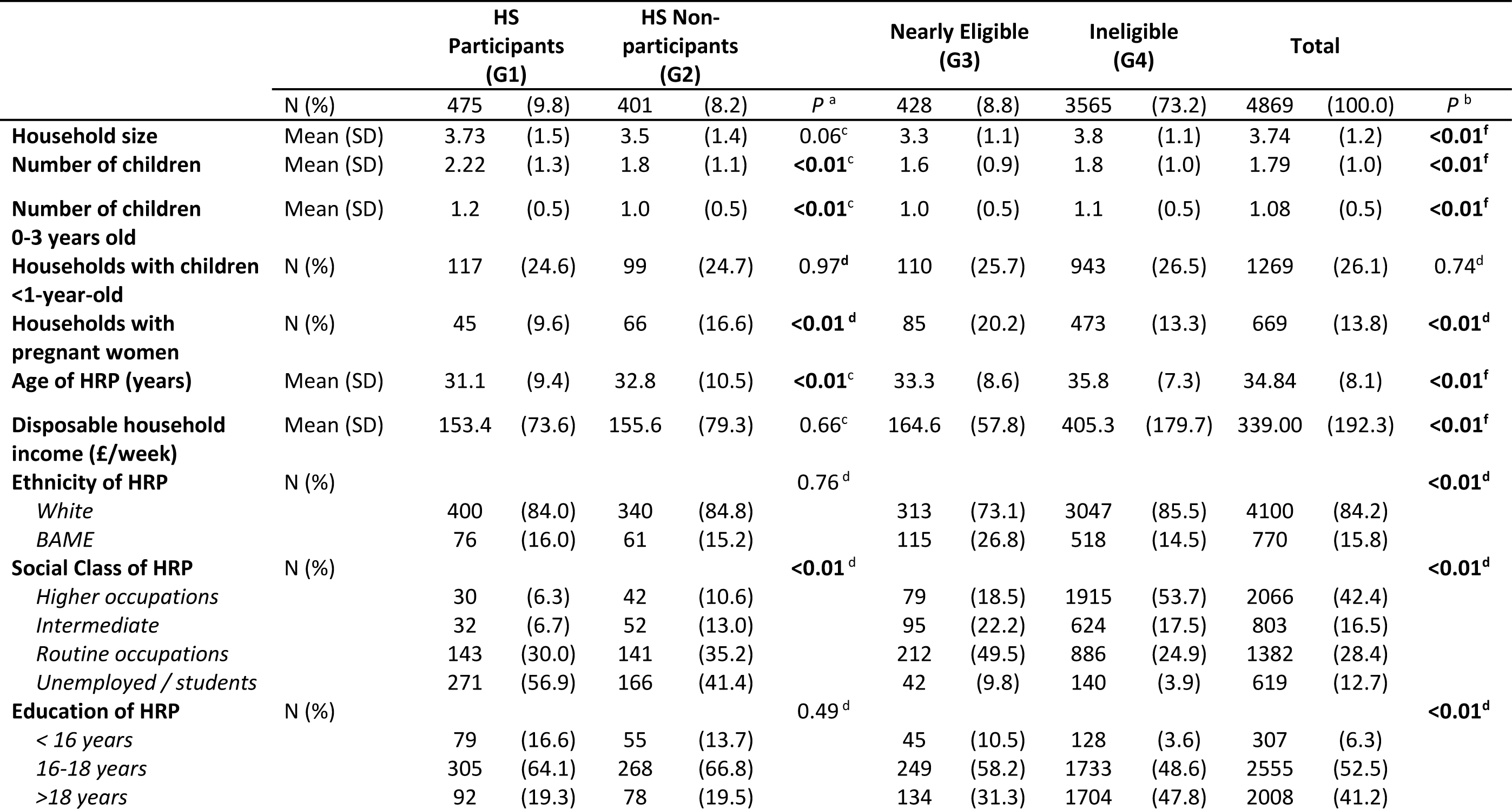
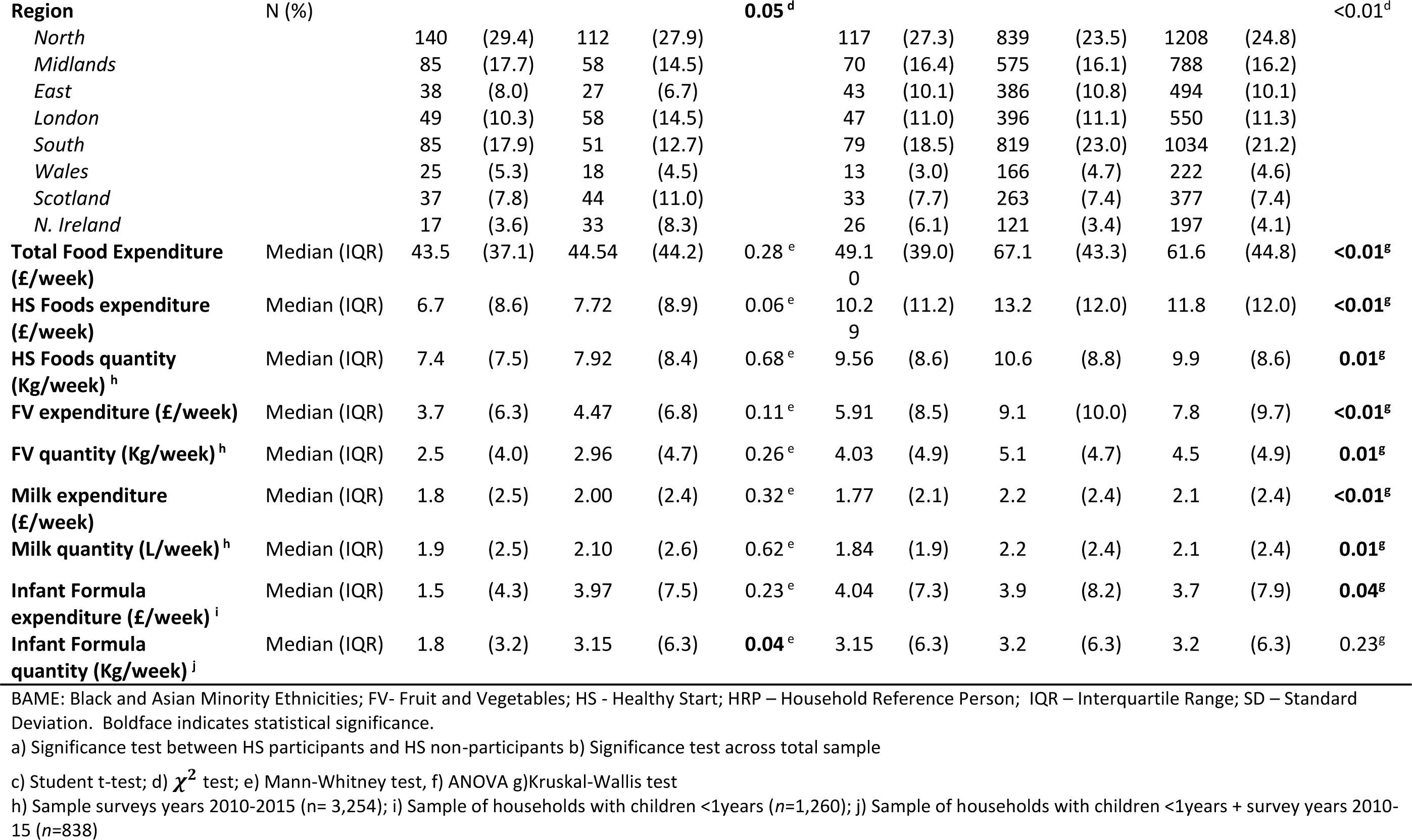
Sample characteristics of households containing children 0-3 years or pregnant women stratified by HS participation

Multivariable quantile regression was used to assess differences in each outcome between the four exposure groups, using Healthy Start Eligible Non-participants (G2) as the reference group. Since outcomes were positively skewed, quantile regression estimates the median (or other percentile) of outcome distribution instead of the mean and is therefore less sensitive to the influence of outliers(18). Quantile regression also allows for the effects of the covariates to differ at different points of the outcome distribution. Results are presented for the 25^th^, 50^th^ and 75^th^ percentiles of the outcome variable.

Ordinary least squares (OLS) regression was also performed and presented alongside results of the quantile regression as a comparison between the two methods and check for robustness. Wald tests were performed to test for equality between Nearly Eligible (G3) and Ineligible (G4) coefficients at the 25^th^, 50^th^ and 75^th^ percentile. Multicollinearity was tested by calculating variation inflation factors (VIF), all values were below 10 (max VIF=1.51) indicating no evidence for multicollinearity.

Covariates were added into the regression models in a stepwise manner. Model 1 adjusted for survey year and survey quarter. Model 2 additionally included household size, number of children and age of HRP. Model 3 additionally included ethnicity of HRP, NS-SEC social class, age HRP completed full-time education and region.

For sensitivity analyses, the same descriptive analyses and quantile regressions on expenditure outcomes were performed after excluding participants without quantity of food purchases data (2015-2017).

Stata V.15 (StataCorp) was used to perform all descriptive and inference tests, using a 95% confidence level for significance.

## RESULTS

Table 1 presents the characteristics of the analytic sample. A total of 876 households were eligible for Healthy Start, of these, 54% (*n*=475) reported participating in Healthy Start and 46% (*n=*401) households were Eligible Non-participants. Healthy Start Participants (G1) and Eligible Non-participants (G2) had similar mean income level, ethnicity and education, but participants (G1) were more likely to be in a lower social class and have young children but were less likely to have a pregnant woman than Eligible Non-participants (G2). Households which were ineligible for Healthy Start (G3+G4) were found to be older and have a higher occupation, education and income levels than eligible households (G1+G2).

Results of the median quantile regression of FV, Healthy Start foods, infant formula and total food expenditure across the four exposure groups are displayed in Table 2. In the minimally adjusted model, a significant lower purchase of FV and HS foods was observed in Healthy Start Participants (G1) compared to Eligible Non-participants (G2). However, differences did not persist. In the fully adjusted models, there was no statistically significant difference between Healthy Start Participants (G1) and Eligible Non-participants (G2) in FV, Healthy Start food or total food expenditure. Infant formula purchases were significantly lower in Healthy Start Participants (G1) (−1.82 £/week; 95% CI −3.12, −0.51). These results were consistent when quantity variables were analysed (Table S2). Cow’s milk was tested as an outcome but there was no difference in expenditure across all groups (Table 2). Nearly Eligible (G3) and Ineligible households (G4), however, were observed with higher FV and Healthy Start food expenditure than Eligible Non-participants (G2). For total food expenditure, only Ineligible households (G4) had significantly higher spending compared to Eligible Non-participants (7.30 £/week; 95% CI 3.06, 11.53).

**Table 2.** Median regression of HS participation on food expenditure in LCFS (years 2010-2017, *n*=4,870)

|  | **Model 1** |  | **Model 2** |  | **Model 3** |  |
| --- | --- | --- | --- | --- | --- | --- |
|  | Coef. | [95% CI] | Coef. | [95% CI] | Coef. | [95% CI] |
| **FV expenditure (£/week)** |  |  |  |  |  |  |
| *HS Participants (G1)* | **-0.89\*** | [-1.67,-0.10] | -0.25 | [-0.80,0.29] | 0.37 | [-0.37,1.11] |
| *HS Non-participants (G2)* | - | - | - | - | - | - |
| *Nearly Eligible (G3)* | **1.56\*\*** | [0.49,2.63] | **1.40\*\*** | [0.49,2.31] | **1.14\*** | [0.18,2.09] |
| *Ineligible (G4)* | **4.56\*\*\*** | [3.88,5.23] | **3.55\*\*\*** | [2.91,4.18] | **2.22\*\*\*** | [1.57,2.86] |
| **HS food expenditure (£/week)** |  |  |  |  |  |  |
| *HS Participants (G1)* | **-1.14\*** | [-2.27,-0.00] | -0.64 | [-1.48,0.20] | -0.07 | [-0.85,0.71] |
| *HS Non-participants (G2)* | - | - | - | - | - | - |
| *Nearly Eligible (G3)* | **2.05\*\*** | [0.81,3.29] | **1.96\*\*\*** | [0.84,3.09] | **1.60\*\*\*** | [0.79,2.41] |
| *Ineligible (G4)* | **5.11\*\*\*** | [4.26,5.97] | **3.69\*\*\*** | [2.86,4.51] | **2.56\*\*\*** | [1.77,3.35] |
| **Infant formula expenditure (£/week) <sup>a</sup>** |  |  |  |  |  |  |
| *HS Participants (G1)* | **-3.07\*\*\*** | [-4.80,-1.35] | **-2.73\*\*** | [-4.51,-0.94] | **-1.82\*\*** | [-3.12,-0.51] |
| *HS Non-participants (G2)* | - | - | - | - | - | - |
| *Nearly Eligible (G3)* | -0.53 | [-1.76,0.70] | -0.61 | [-1.95,0.73] | -0.54 | [-1.91,0.83] |
| *Ineligible (G4)* | -0.44 | [-1.49,0.61] | -0.35 | [-1.52,0.82] | -0.83 | [-2.04,0.38] |
| **Total food expenditure (£/week)** |  |  |  |  |  |  |
| *HS Participants (G1)* | -0.31 | [-5.99,5.37] | -4.11 | [-9.46,1.25] | -1.39 | [-5.72,2.95] |
| *HS Non-participants (G2)* | - | - | - | - | - | - |
| *Nearly Eligible (G3)* | 4.52 | [-0.02,9.06] | 1.61 | [-3.97,7.19] | 2.65 | [-2.19,7.48] |
| *Ineligible (G4)* | **21.85\*\*\*** | [17.58,26.13] | **13.43\*\*\*** | [8.69,18.18] | **7.30\*\*\*** | [3.06,11.53] |
| **Cow's milk expenditure (£/week)** |  |  |  |  |  |  |
| *HS Participants (G1)* | -0.24 | [-0.53,0.04] | -0.25 | [-0.53,0.02] | -0.17 | [-0.43,0.09] |
| *HS Non-participants (G2)* | - | - | - | - | - | - |
| *Nearly Eligible (G3)* | -0.2 | [-0.49,0.08] | -0.15 | [-0.35,0.06] | -0.21 | [-0.46,0.03] |
| *Ineligible (G4)* | 0.14 | [-0.11,0.39] | -0.02 | [-0.22,0.18] | -0.18 | [-0.41,0.05] |

LCFS – Living Costs and Food Survey; CI – confidence interval; FV – Fruit and Vegetables; HS - Healthy Start; HRP – Household Reference Person
Boldface indicates statistical significance \* $P<0.05$ \*\* $P<0.01$ \*\*\* $P<0.001$
a) Sample of households with children <1years ( $n=1,260$ )
Model 1 – Adjusted for year + quarter
Model 2 – Adjusted for Model 1, household size, number of children <1 year, 0-3 years + age of HRP
Model 3 – Adjusted for Model 2, region, ethnicity, social class and education of HRP

We additionally assessed the differences in outcome at the 25^th^ and 75^th^ percentile using quantile regression across the four exposure groups. This is important as the difference in spending between Healthy Start Eligible (G1+G2), Nearly Eligible (G3) and Ineligible (G4) households differed across the expenditure distribution. For example, the non-significant differences in FV expenditure between Health Start Participants (G1) and Eligible Non-participants (G2) were observed consistently at the 25^th^, 50^th^ and 75^th^ percentile (Figure 1.A). However, differences in FV expenditure of Nearly Eligible (G3) and Ineligible (G4) compared to Healthy Start Eligible Non-participants (G2) increased between the 25^th^ and 75^th^ percentile of FV expenditure (Figure 1.A). This implies that the more ineligible households (G3+G4) spent on FV, the greater the magnitude of difference compared to Healthy Start Eligible Non-participating households (G2). Importantly, a similar pattern was not seen for FV quantity. The coefficients were of consistent magnitude across all percentiles assessed (Figure 1.B). This indicates that the higher levels of expenditure observed did not correspond to a higher quantity of FV purchased.

**Figure 1.**
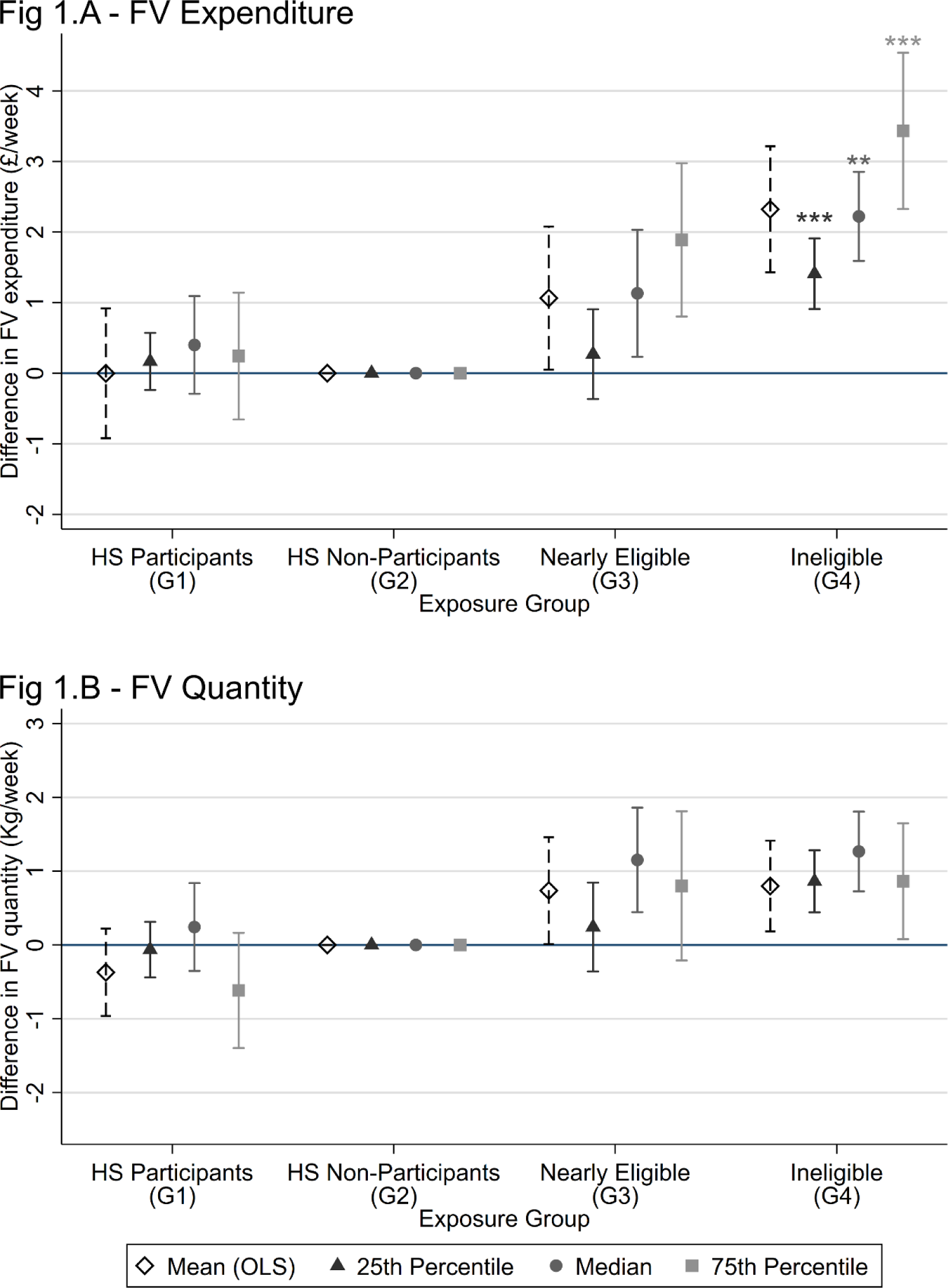
Quantile regression of FV expenditure and quantity by Healthy Start participation. Footnotes: Significant difference between nearly eligible and ineligible groups using a Wald test *P <0.05, **P <0.01 *** P <0.001. FV- Fruit and vegetables; HS- Healthy Start; OLS- Ordinary Least Squares regression. A) FV expenditure (£/week) years 2010-17, n=4,870; B) FV quantity (Kg/week), years 2010-15, n=3,265.

Sensitivity analyses demonstrated that results were robust when using a complete case analysis (Table S2 and S3).

## DISCUSSION

Using nationally representative data, the present analysis did not find evidence of an association between Healthy Start participation and the purchase of FV, Healthy Start foods or total food. An inequality in purchases was observed as FV expenditure was higher in both Nearly Eligible (G3) and Ineligible households (G4), compared to eligible households (G1+G2). Total food expenditure was higher only in Ineligible households (G4).

No previous evaluation of the scheme has compared the impact of the Healthy Start programme within an eligible population. Griffith et al(11) used a difference-in-differences analysis of household purchase data two years before and after programme implementation, reporting a £2.43/month (£0.61/week) increase in FV spending in Healthy Start eligible households compared to ineligible low-income households with a child aged 4-8 years. Scantlebury et al(10) compared FV intake among adults and children aged 5 years or over from Healthy Start eligible and ineligible households in England, but reported no association between Healthy Start eligibility and individual FV intake following programme introduction. The present finding adds to the current evidence base, indicating that it is unlikely that Healthy Start vouchers had a discernible impact on the dietary behaviours of its target population. In lieu of an experimental design, our study has used the most appropriate control group, eligible non-participants, to evaluate the effect of the voucher.

By contrast, a similar food assistance programme in the United States has demonstrated greater success. The Special Supplemental Nutrition Program for Women, Infants and Children (WIC) serves low-income families with pregnant women or children aged 0-5 years at risk of nutritional deficiencies. However, alongside distributing cash-value vouchers for FV, WIC additionally provides healthy food packages. Despite some inconsistencies(19, 20), evaluations of WIC report improved dietary intake(21–23) and infant health outcomes(24) in WIC participants compared to eligible non-participants. The larger total benefit received by WIC participants compared to Healthy Start could explain why the programme appears more successful.

We did not find evidence of an association between Healthy Start participation and FV purchases. Economic theory suggests that only households which previously spent less than the voucher value on target foods will increase their spending on these items(11, 25). Otherwise, the voucher will act as financial assistance, permitting money in the budget to be spent elsewhere. The Healthy Start voucher has not changed value since 2010, as such price inflation over this period has undermined the voucher value. Consequently, it is unlikely that the voucher provided enough purchasing power to increase FV expenditure above usual levels in low-income households. As FV prices in the UK are set to rise(26), it is concerning that the voucher may have a further diminished value in the future. In response to this issue, the Scottish government raised the weekly value of the Healthy Start voucher to £4.25 in August 2019(27). In this way, increasing the benefit may enable Healthy Start participants to make a more meaningful change to their diet.

Health professionals have expressed concern that the inclusion of infant formula in the Healthy Start scheme may discourage breastfeeding(28). In this analysis, Healthy Start Participants (G1) purchased a significantly lower amount of infant formula compared to Eligible Non-participants (G2), which could neither be explained by differences in total food expenditure nor differing prevalence of infants in the households. However, breastfeeding rates were unobserved thus could not be controlled for in this analysis. Findings from a Scottish longitudinal cohort suggest infant feeding practices were not significantly different between Healthy Start recipients and other nearly eligible mothers(29). Together, these results could suggest Healthy Start does not disincentivise breast-feeding, however further investigation is needed to confirm this hypothesis.

An inequality in FV purchases was apparent between relatively low-income and higher-income households, reinforcing previous literature that income is associated with FV purchasing behaviours(1, 3). A higher quantity of FV purchased in Nearly Eligible households (G3) compared to eligible households (G1+G2) indicates that the programme may not mitigate even small income-inequalities.

This study was novel in its ability to characterise Healthy Start Participants (G1) compared to Eligible Non-participants (G2). We found that households with pregnant women were less likely to participate in the Healthy Start scheme. This is supported by qualitative research reporting poor awareness of the scheme amongst pregnant women(30). A reliance on health professionals to promote the scheme has meant eligible pregnant women frequently learnt of the programme after birth. Improving universal awareness of the scheme may increase uptake(9). Moreover, in April 2020 the requirement for a health professional’s signature on application forms was removed (Table S1), possibly improving future uptake of the scheme.

### Strengths and limitations

The strengths of the present study are that it is the first to use a nationally representative dataset for the UK and observed participation in the Healthy Start scheme. We were able to accurately define a range of exposure groups due to the use of data containing detailed variables on household composition and income. Our results were also robust to a range of sensitivity analyses on the potential impacts of missing data.

The primary limitation is that the data were cross-sectional, therefore change in participant’s purchasing behaviours as a result of the vouchers could not be determined. Additionally, pooling years limited the ability to account for macroeconomic changes over time, although we did adjust for inflation and include year and quarter indicators in analyses to reduce potential biases. Additionally, as Healthy Start is targeted at very low-income households, the number of eligible households in nationally representative data was low. Resultantly, the analysis was underpowered to determine significance in small differences of FV expenditure. Finally, although participation in the scheme was self-reported, all reported incomes were confirmed with documentation, minimizing any potential misclassification bias.

## Conclusion

In summary, our analysis did not provide evidence of different FV, Healthy Start foods or total food expenditure between Healthy Start participants and non-participants. The observed socioeconomic gradient in food spending reflects continuing inequalities in the UK. Our findings implicate that actions to improve Healthy Start, such as increasing voucher value and improving uptake, would be needed for the programme to better serve the target population. This evaluation provides valuable shared lessons for similar food-assistance programs worldwide; iterative evaluations of food assistance programs are needed to ensure they dynamically meet the needs of low-income families.

## Data Availability

The datasets generated and/or analysed during the current study are available in the UK data service (GN 33334).

http://doi.org/10.5255/UKDA-SN-8459-2

## Abbreviations

UK: United Kingdom
FV: Fruit and vegetables
LCFS: Living Costs and Food Survey
OECD: Organisation for Economic Co-operation and Development
HRP: Household reference person
BAME: Black and Asian minority ethnicities
NS-SEC: National Statistics Socio-economic Classification
WIC: Special Supplemental Nutrition Program for Women, Infants and Children
G1: Group 1
G2: Group 2
G3: Group 3
G4: Group 4
OLS: Ordinary Least Squares
VIF: Variance inflation factor
HS: Healthy Start
SD: Standard deviation
CI: Confidence interval
IQR: Interquartile Range
ANOVA: Analysis of variance

## DECLARATIONS

### Ethics Approval

Imperial College Research Ethics Committee (ICREC) have confirmed that ethical approval was not required for this study. This study was a secondary analysis of a dataset accessed via the UK data service.

### Availability of data and materials

The datasets generated and/or analysed during the current study are available in the UK data service (GN 33334). Accessed from http://doi.org/10.5255/UKDA-SN-8459-2

### Author contributions

EPV, JPS, SvH and CM conceptualised the study. JCP, AL and EPV designed the study. SvH and KC advised on methods. JCP ran statistical analyses. All authors contributed to writing and approved final article.

## Acknowledgements

The NIHR School for Public Health Research is a partnership between the Universities of Sheffield; Bristol; Cambridge; Imperial College London; and University College London; The London School for Hygiene and Tropical Medicine (LSHTM); LiLaC – a collaboration between the Universities of Liverpool and Lancaster; and Fuse - The Centre for Translational Research in Public Health a collaboration between Newcastle, Durham, Northumbria, Sunderland and Teesside Universities

