## Supplementary material for "Is the Healthy Start scheme associated with increased food expenditure in low-income families with young children in the United Kingdom?": Table S1

**Supplemental Table 1 -** Healthy Start programme eligibility criteria and provision[29,30]

| Beneficiary | Eligibility requirement | Entitlement | List of items accepted |
|---|---|---|---|
| **Pregnant woman aged <18 years** | No requirement. | £3.10/week<br>(USD 16/month) | **Fresh or frozen fruit and vegetables**<br>- not including fruit or vegetables to which fat, salt, sugar, flavoring or any other ingredients have been added<br>**Liquid cow's milk**<br>- not including milk to or from which chemicals, vitamins, flavors or colors have been added or removed<br>**Infant formula**<br>- From birth – one year, cow's milk formula |
| **Pregnant woman aged 18+ years** | (i) Someone in household is entitled to:<br>- income support<br>- an income-based jobseeker's allowance<br>- universal credit and has earned income of £408 or less<br>- child tax credit and household income below £16,190*<br><br>and (ii) Application approved by health professional (before April 2020) | £3.10/ week<br>(USD 16/month) | |
| **A child aged < one year** | | £6.20/ week<br>(USD 32/month) | |
| **A child aged 1-3 years** | | £3.10/ week<br>(USD 16/month) | |

*Tax credit threshold: 2005 - £13,190; 2006 - £14,155; 2007 - £14,495; 2008 - £15,575; 2009 - £16,040; 2010-19 - £16,190

**Supplemental Table 2**– Sample characteristics of households containing children 0-3 years or pregnant women in the Living Costs and Food survey, UK, (years 2010-15) stratified by HS participation.

| | | **HS participants** | | **HS non-participants** | | | **Nearly Eligible** | | **Ineligible** | | **Total** | | |
|---|---|---|---|---|---|---|---|---|---|---|---|---|---|
| | N (%) | 344 | (10.57) | 281 | (8.64) | *P** | 267 | (8.21) | 2362 | (72.59) | 3254 | (100) | *P*† |
| **Household size** | Mean (SD) | 3.69 | (1.55) | 3.52 | (1.35) | 0.14‡ | 3.24 | (1.00) | 3.79 | (1.09) | 3.71 | (1.17) | <0.01# |
| **Number of children** | Mean (SD) | 2.19 | (1.33) | 1.82 | (1.09) | <0.01‡ | 1.44 | (0.86) | 1.74 | (1.01) | 1.77 | (1.06) | <0.01# |
| **Number of children 0-3 years old** | Mean (SD) | 0.24 | (0.44) | 0.23 | (0.43) | <0.01‡ | 0.26 | (0.44) | 0.27 | (0.45) | 0.26 | (0.45) | 0.<0.01# |
| **Households with children <1 year old** | N (%) | 81 | (23.55) | 64 | (22.78) | 0.82§ | 70 | (26.22) | 623 | (26.38) | 838 | (25.75) | 0.44§ |
| **Households with pregnant women** | N (%) | 37 | (10.95) | 48 | (17.27) | 0.02§ | 53 | (20.23) | 330 | (14.04) | 468 | (14.49) | <0.01§ |
| **Age of HRP (years)** | Mean (SD) | 30.16 | (9.00) | 32.81 | (10.33) | <0.01‡ | 32.82 | (8.61) | 35.61 | (7.29) | 34.56 | (8.11) | <0.01# |
| **Equivalised gross household income (£/week)** | Mean (SD) | 158.69 | (82.59) | 163.16 | (87.86) | 0.51‡ | 176.75 | (62.83) | 479.38 | (297.18) | 393.34 | (292.29) | <0.01# |
| **Equivalised disposable household income (£/week)** | Mean (SD) | 146.74 | (72.84) | 151.83 | (76.21) | 0.40‡ | 160.12 | (57.23) | 393.98 | (174.68) | 327.74 | (187.36) | <0.01# |
| **Ethnicity of HRP** | N (%) | | | | | 0.59§ | | | | | | | <0.01§ |
| *White* | | 291 | (84.59) | 242 | (86.12) | | 199 | (74.53) | 2033 | (86.07) | 2765 | (84.97) | |
| *BAME* | | 53 | (15.41) | 39 | (13.88) | | 68 | (25.47) | 329 | (13.93) | 489 | (15.03) | |
| **Social Class of HRP** | N (%) | | | | | <0.01§ | | | | | | | <0.01§ |
| *Higher managerial occupations* | | 18 | (5.23) | 25 | (8.90) | | 44 | (16.48) | 1239 | (52.46) | 1326 | (40.75) | |
| *Intermediate occupations* | | 18 | (5.23) | 33 | (11.74) | | 67 | (25.09) | 416 | (17.61) | 534 | (16.41) | |
| *Routine and manual occupations* | | 91 | (26.45) | 96 | (34.16) | | 127 | (47.57) | 605 | (25.61) | 919 | (28.24) | |
| *Unemployed or students* | | 217 | (63.08) | 127 | (45.20) | | 29 | (10.86) | 102 | (4.32) | 475 | (14.60) | |
| **Education of HRP** | N (%) | | | | | 0.82§ | | | | | | | <0.01§ |
| < 16 years | | 57 | (16.57) | 42 | (14.95) | | 28 | (10.49) | 88 | (3.73) | 215 | (6.61) | |
| 16 – 18 years | | 225 | (65.41) | 190 | (67.62) | | 150 | (56.18) | 1182 | (50.04) | 1747 | (53.69) | |
| >18 years | | 62 | (18.02) | 49 | (17.44) | | 89 | (33.33) | 1092 | (46.23) | 1292 | (39.70) | |

| | | | | | | | | | | | | | |
|---|---|---|---|---|---|---|---|---|---|---|---|---|---|
| **Region** | N (%) | | | | | <0.01§ | | | | | | | <0.01§ |
| *North* | | 109 | (31.69) | 87 | (30.96) | | 75 | (28.09) | 550 | (23.29) | 821 | (25.23) | |
| *Midlands* | | 63 | (18.31) | 39 | (13.88) | | 47 | (17.60) | 394 | (16.68) | 543 | (16.69) | |
| *East* | | 31 | (9.01) | 13 | (4.63) | | 21 | (7.87) | 245 | (10.37) | 310 | (9.53) | |
| *London* | | 34 | (9.88) | 41 | (14.59) | | 33 | (12.36) | 268 | (11.35) | 376 | (11.56) | |
| *South* | | 59 | (17.15) | 35 | (12.46) | | 52 | (19.48) | 561 | (23.75) | 707 | (21.73) | |
| *Wales* | | 18 | (5.23) | 15 | (5.34) | | 11 | (4.12) | 111 | (4.70) | 155 | (4.76) | |
| *Scotland* | | 25 | (7.27) | 27 | (9.61) | | 17 | (6.37) | 172 | (7.28) | 241 | (7.41) | |
| *N. Ireland* | | 5 | (1.45) | 24 | (8.54) | | 11 | (4.12) | 61 | (2.58) | 101 | (3.10) | |
| **Total Food Expenditure (£/week)** | Median (IQR) | 42.60 | (35.41) | 42.57 | (41.23) | 0.44¶ | 46.77 | (39.07) | 66.72 | (43.13) | 60.74 | (44.36) | <0.01\|\| |
| **Total HS Foods expenditure (£/week)** | Median (IQR) | 6.73 | (8.21) | 7.61 | (8.30) | 0.12¶ | 9.91 | (11.07) | 13.03 | (11.60) | 11.54 | (11.65) | <0.01\|\| |
| **Total HS Foods quantity (Kg/week)** | Median (IQR) | 7.41 | (7.51) | 7.92 | (8.37) | 0.68¶ | 9.56 | (8.57) | 10.56 | (8.84) | 9.89 | (8.62) | <0.01\|\| |
| **FV expenditure (£/week)** | Median (IQR) | 3.33 | (5.92) | 4.12 | (6.65) | 0.12¶ | 5.77 | (7.90) | 9.00 | (9.60) | 7.64 | (9.38) | <0.01\|\| |
| **FV quantity (kg/week)** | Median (IQR) | 2.46 | (3.97) | 2.96 | (4.69) | 0.26¶ | 4.03 | (4.92) | 5.05 | (4.73) | 4.51 | (4.89) | <0.01\|\| |
| **Cow's milk expenditure (L/week)** | Median (IQR) | 1.85 | (2.46) | 2.10 | (2.57) | 0.62¶ | 1.84 | (1.92) | 2.23 | (2.44) | 2.14 | (2.43) | <0.01\|\| |
| **Infant Formula expenditure (£/week) ††** | Median (IQR) | 1.90 | (4.04) | 3.82 | (7.41) | 0.03¶ | 3.83 | (7.27) | 3.80 | (8.03) | 3.72 | (7.55) | 0.13\|\| |
| **Infant Formula expenditure (Kg/week) ††** | Median (IQR) | 1.75 | (3.15) | 3.15 | (6.30) | 0.04¶ | 3.15 | (6.30) | 3.15 | (6.30) | 3.15 | (6.30) | 0.23\|\| |

Note: SD – Standard Deviation; IQR – Interquartile Range; HS - Healthy Start; HRP – Household Reference Person; BAME: Black and Minority Ethnicities; FV – Fruit and Vegetables

* Significance difference between HS participants and HS non-participants † Significance difference across total sample

‡ Student t-test; § X2 test ; ¶ Mann-Whitney test; # ANOVA; ||Kruskal-Wallis test

†† Sample of households with children <1years + survey years 2010-15 (n=838)

**Supplemental Table 3 -** Quantile regression of HS participation on food expenditure and quantity in the Living Costs and Food survey, UK, years 2010-2015 (*n*=3,254)

| | **Model 1** | | **Model 2** | | **Model 3** | |
|---|---|---|---|---|---|---|
| | Coef. | [95% CI] | Coef. | [95% CI] | Coef. | [95% CI] |
| **FV expenditure (£/week)** | | | | | | |
| *HS participants* | -1.10* | [-1.96,-0.23] | 0.14 | [-0.59,0.87] | 0.44 | [-0.26,1.14] |
| *HS non-participants* | - | - | - | - | - | - |
| *Nearly Eligible* | 1.84** | [0.61,3.07] | 2.07*** | [0.91,3.22] | 1.64** | [0.57,2.71] |
| *Ineligible* | 4.50*** | [3.79,5.21] | 3.84*** | [3.10,4.58] | 2.36*** | [1.65,3.08] |
| **FV quantity (Kg/week)** | | | | | | |
| *HS participants* | -0.35 | [-1.09,0.39] | 0.1 | [-0.47,0.67] | 0.23 | [-0.32,0.79] |
| *HS non-participants* | - | - | - | - | - | - |
| *Nearly Eligible* | 1.31*** | [0.55,2.07] | 1.29*** | [0.67,1.91] | 1.05** | [0.27,1.83] |
| *Ineligible* | 2.24*** | [1.57,2.91] | 1.61*** | [1.15,2.06] | 1.06*** | [0.52,1.60] |
| **HS food expenditure (£/week)** | | | | | | |
| *HS participants* | -1.14 | [-2.35,0.07] | -0.47 | [-1.60,0.66] | 0.09 | [-0.67,0.85] |
| *HS non-participants* | - | - | - | - | - | - |
| *Nearly Eligible* | 2.14** | [0.78,3.50] | 2.38*** | [0.98,3.78] | 2.42*** | [1.35,3.50] |
| *Ineligible* | 4.96*** | [4.01,5.90] | 3.83*** | [2.80,4.87] | 2.72*** | [1.86,3.59] |
| **HS food quantity (Kg/week)** | | | | | | |
| *HS participants* | -0.63 | [-1.66,0.40] | -0.24 | [-1.28,0.80] | -0.51 | [-1.56,0.53] |
| *HS non-participants* | - | - | - | - | - | - |
| *Nearly Eligible* | 1.18 | [-0.17,2.54] | 1.65** | [0.55,2.74] | 1.30* | [0.13,2.47] |
| *Ineligible* | 2.37*** | [1.42,3.32] | 1.40** | [0.43,2.37] | 0.92 | [-0.15,2.00] |
| **Infant formula expenditure (£/week)** † | | | | | | |
| *HS participants* | -2.73** | [-4.50,-0.96] | -2.87** | [-4.59,-1.16] | -2.45*** | [-3.67,-1.23] |
| *HS non-participants* | - | - | - | - | - | - |
| *Nearly Eligible* | -0.72 | [-2.55,1.11] | -0.9 | [-2.63,0.84] | -1.03 | [-2.67,0.62] |
| *Ineligible* | -0.58 | [-1.96,0.79] | -0.74 | [-2.10,0.62] | -1.73* | [-3.20,-0.26] |
| **Infant formula quantity (Kg/week)** † | | | | | | |
| *HS participants* | -1.4 | [-2.93,0.13] | -1.38 | [-3.01,0.24] | -1.58* | [-3.10,-0.06] |
| *HS non-participants* | - | - | - | - | - | - |
| *Nearly Eligible* | 0 | [-1.25,1.25] | -0.06 | [-1.47,1.35] | -1.14 | [-2.96,0.67] |
| *Ineligible* | 0 | [-0.63,0.63] | -0.08 | [-0.73,0.58] | -0.85 | [-1.87,0.16] |

**P*<0.05 ***P*<0.01 ****P*<0.001*

† Sample of households with children <1years (*n*=838)

Model 1 – Adjusted for year + quarter

Model 2 – Adjusted for Model 1, household size, number of children <1 year, 0-3 years + age of HRP

Model 3 – Adjusted for Model 2, region, ethnicity, social class and education of HRP
